# Systematic Review and Meta-Analysis of Endovascular Therapy Effectiveness for Unruptured Saccular Intracranial Aneurysms

**DOI:** 10.1101/2023.07.12.23292485

**Authors:** Sergio A. Pineda-Castillo, Evan R. Jones, Keely A. Laurence, Lauren R. Thoendel, Tanner L. Cabaniss, Yan D. Zhao, Bradley N. Bohnstedt, Chung-Hao Lee

## Abstract

**Background:** Currently, endovascular treatment of intracranial aneurysms (ICAs) is limited by low complete occlusion rates. The advent of novel endovascular technology has expanded the applicability of endovascular therapy; however, the superiority of novel embolic devices over the traditional Guglielmi detachable coils (GDCs) is still debated. We performed a systematic review of literature that reported the Raymond-Roy occlusion classifications (RROC) rates of modern endovascular devices to determine their immediate and long-term occlusion effectiveness for the treatment of unruptured saccular ICAs.

**Methods:** A search was conducted using electronic databases (PUBMED, Cochrane, ClinicalTrials.gov, Web of Science). We retrieved studies published between 2000-2022 reporting immediate and long-term RROC rates of subjects treated with different endovascular ICA therapies. We extracted demographic information of the treated patients and their reported angiographic RROC rates.

**Results:** A total of 80 studies from 15 countries were included for data extraction. RROC rates determined from angiogram were obtained for 21,331 patients (72.5% females, pooled mean age: 58.2 (95% CI: 56.8-59.6), harboring 22,791 aneurysms. The most frequent aneurysm locations were the internal carotid artery (46.4%, 95% CI: 41.9%-50.9%), the anterior communicating artery (26.4%, 95% CI: 22.5%-30.8%), the middle cerebral artery (24.5%, 95% CI:19.2%-30.8%) and the basilar tip (14.4%, 95% CI:11.3%-18.3%). The RROC rates were analyzed for GDCs, the Woven EndoBridge (WEB), and flow diverters. The immediate complete occlusion (RROC-I) rate was the highest in balloon-assisted coiling (73.9%, 95% CI: 65.0%-81.2%) and the lowest in the WEB (27.8%, 95% CI:13.2%-49.2%). The long-term complete occlusion probability was homogenous in all analyzed devices.

**Conclusions:** We observed that the coil-based endovascular therapy provides acceptable rates of complete occlusion, where the balloon-assisted coils provide greater probability of immediate complete occlusion. Out of the analyzed devices, the WEB exhibited the shortest time to achieve >90% probability of long-term complete occlusion (∼18 months). Overall, the GDCs remain the *gold standard* for endovascular treatment of unruptured saccular aneurysms.

## INTRODUCTION

Endovascular treatment of intracranial aneurysms (ICA) emerged in 1991 with the invention of the Guglielmi detachable coils (GDCs).^1,2^ Ever since, endovascular treatment has become the preferred method for ICA therapy due to its minimal invasiveness, lower in-hospital fatalities, and superior clinical outcomes, when compared to the traditional, highly invasive surgical clipping.^3-6^ In addition, the coupling of GDCs and other assisting technology, such as balloons and stents, has increased their range of applications and effectiveness, especially for aneurysm with a complex 3D geometry or a high neck-to-sack ratio.^7^

In the last two decades, novel endovascular devices have been designed to expand the application of this minimally invasive method for ICA therapy. The most prominent inventions include bioactive and hydrogel-coated GDCs and flow diverters.^7,8^ For flow diverters, numerous devices have emerged in two categories: (i) extra-saccular: including the p64,^9^ the Pipeline,^10^ the Flow Redirection Endoluminal Device (FRED),^11^ the Surpass,^12^ among others; and (ii) intra-saccular flow diverters: such as the Woven EndoBridge (WEB)^13^ and the Contour.^14^ The advance of this technology has evolved the application of endovascular therapy for complex aneurysm geometries and even aneurysm locations within the neuro-vasculature.

Despite these advancements, current literature has shown that the modern endovascular devices do not demonstrate superior complete occlusion rates than the traditional GDCs for the treatment of unruptured ICAs.^15^ Considering that incomplete occlusion drives the recurrence of the treated aneurysm,^16^ it is of paramount importance that advances in endovascular therapy are aimed at improving the limited complete occlusion rates of the GDCs. However, meta-analyses aimed to compare the occlusion effectiveness of different endovascular devices are limited by a low sample size (i.e., low number of analyzed studies) and the use of few timepoints for the comparison of long-term occlusion effectiveness. Owing to the great success of novel technologies for endovascular therapy, in this study our objective is to perform a comprehensive assessment of the complete occlusion effectiveness of modern endovascular devices to re-evaluate the notion of the GDCs as the *gold standard* in endovascular therapy of unruptured saccular ICAs. In addition, we also present a long-term occlusion effectiveness analysis of modern endovascular devices.

## METHODS

### Literature Search

A systematic literature search was conducted by following the recommendations of Tawfik *et al*., ^17^ the Preferred Reporting Items for the Systematic Reviews and Meta-Analyses (PRISMA) statement,^18^ and the Cochrane Handbook for Systematic Reviews of Interventions.^19^ Briefly, a search was performed using online databases available at our institution: PubMed, Cochrane, Web of Science, and ClinicalTrials.gov. The search was conducted to obtain both prospective and retrospective studies that were conducted to evaluate the occlusion efficacy of endovascular devices for treating unruptured saccular ICAs, including GDCs, hydrogel-coated coils, WEB, flow diverters, liquid embolic devices, and others. Only peer-reviewed studies published between 2000-2022, with recruitment start date on or after 1999, were included in the screening process. A complete set of queries for the database search can be found in **Supplemental Data S1**.

### Inclusion/Exclusion of Studies

Screening of the search results was performed using a list of Inclusion/Exclusion (I/E) criteria; individual studies not adhering to the I/E criteria were rejected for data collection. Some key inclusion criteria (IC) were:

1. Reporting of the occlusion rates with Raymond-Roy occlusion classification (RROC) or a translatable scale.
2. Treating unruptured saccular aneurysms only (in the case of including ruptured aneurysms in study design, occlusion rates must be reported separately per rupture status).
3. Reporting of long-term follow-up of ICA occlusion.
4. Endovascular treatment must be performed as the first line of treatment for the target ICA.

Exclusion criteria (EC) included:

1. Literature reviews.
2. Case studies.
3. Unrelated articles.
4. Animal studies.
5. Studies including trauma-induced, mycotic, pathogen-induced aneurysms.
6. Studies that reported occlusion rates with no discrimination of saccular and non-saccular geometries (i.e., saccular geometries should be reported separately).

A complete list of the I/E criteria can be found in **Supplemental Data S2**. The first stage of screening was performed by three independent reviewers using a spreadsheet containing the title and abstract of the search results after removing duplicates. Then, the second screening was performed by four reviewers by reading the full manuscript. All the studies included after screening were used for data collection.

### Data Collection

Data collection was performed by two reviewers. Studies were distributed proportionally between reviewers, and data was extracted using a standardized spreadsheet that was previously pilot tested. Data extracted for analysis included: (i) *basic study information* (first author, country, publication date, sample size), (ii) *patient demographics* (age and sex), (iii) *aneurysm dimensions* (largest diameter and neck length), (iv) *location in the Circle of Willis* (**Table 1**), and (v) *aneurysm occlusion effectiveness results* (type of endovascular device, angiographic assessment time, follow-up time, number of aneurysms assessed, and RROC rates). After initial data collection, studies were re-distributed among the reviewers to perform another step of data verification, to confirm correct data extraction.

**Table 1.**
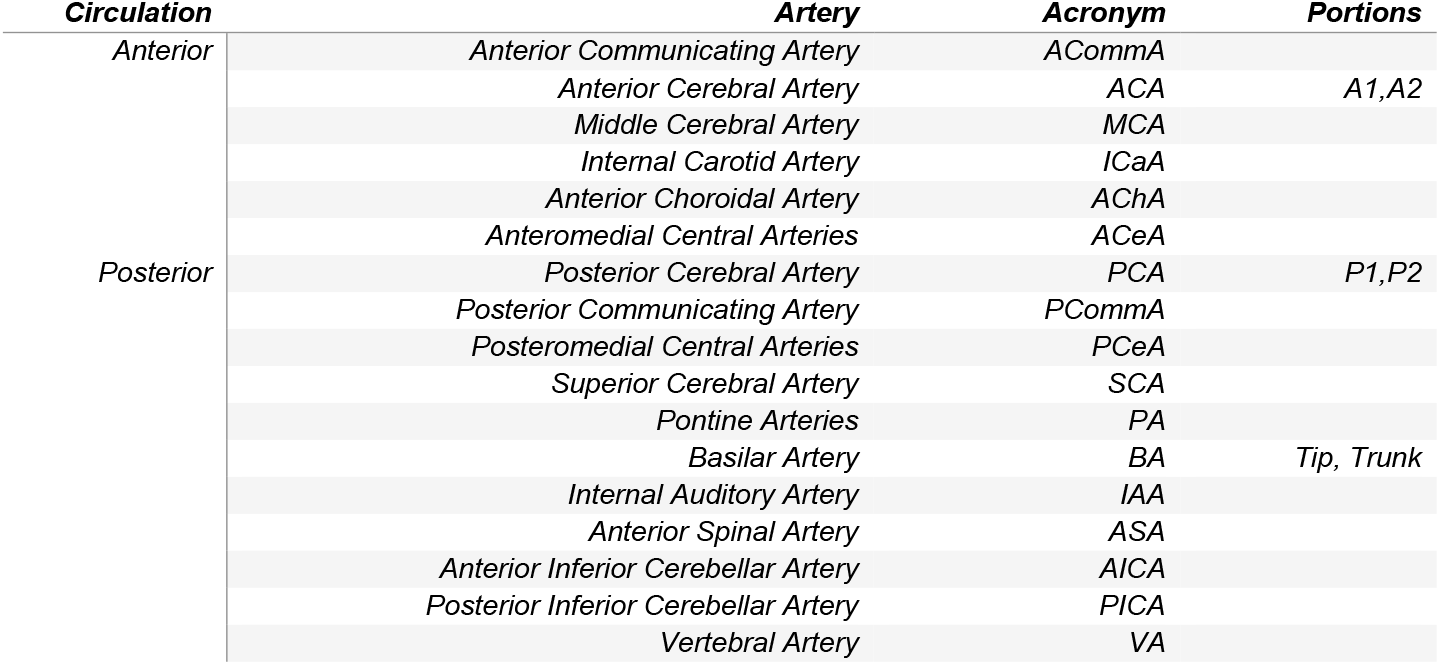
List of aneurysm locations at the Circle of Willis and their corresponding acronyms.

### Outcomes

The objective of this meta-analysis was to compare the immediate and long-term occlusion rates of modern endovascular devices for treating unruptured ICAs. In addition, the demographic information about ICA size and location on the Circle of Willis was collected and the corresponding meta-averages and meta-proportions were analyzed.

### Statistical Analysis

Summary statistics reported in individual studies were transformed to obtain the mean ± standard deviation (SD) of each extracted metric. Proportions, averages, and their respective 95% confidence interval (CI) were pooled across studies using a random-effects mixed model meta-analysis. Heterogeneity was quantified using *I*^2^. A logistic generalized linear mixed model (LGLMM) was used to compare the time-dependent complete occlusion probability between different endovascular devices. All statistical analyses were performed in R v4.1.2 using the meta-package ^20^.

## RESULTS

### Literature Search Results and Screening

Literature search was performed on November 7^th^, 2022, and yielded 5,227 results, which were included in the screening pipeline presented in **Figure 1**. A total of 80 studies were included for data extraction. A complete list of the included studies is provided in **Supplemental Data 3**.

**Figure 1.**
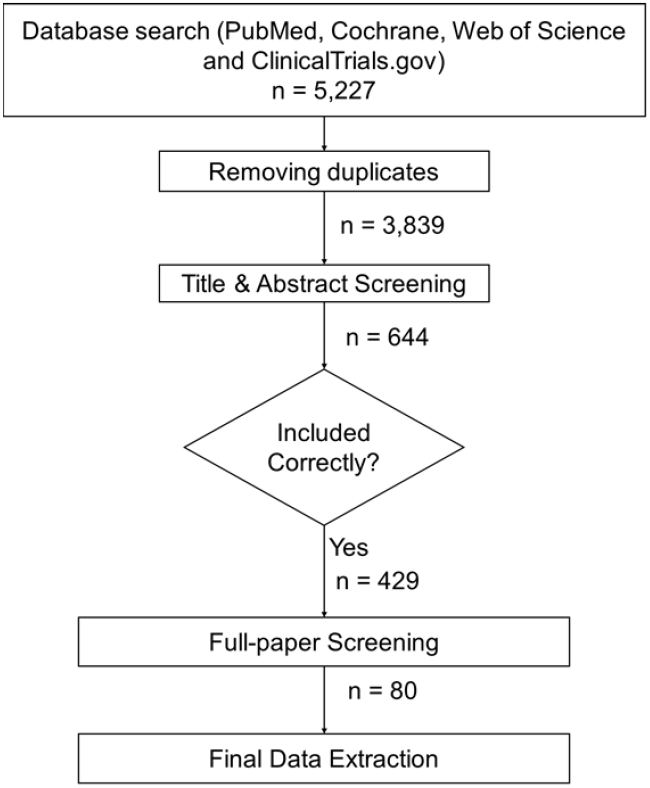
Flow diagram of the screening of obtained studies. *n* represents the number of studies that were included after each phase of the screening process.

### Patient and Study Demographics

In total, the 80 studies used for data extraction included 21,331 patients from 15 countries that harbored 22,791 aneurysms. See **Supplemental Data S4** for these studies included in data extraction. The pooled female proportion was 72.5% (95% CI: 70.9%-74.2%). The pooled mean patient age was 58.2 (95% CI: 56.8-59.6, **Table 2**). The overall aneurysm maximum diameter average and neck size were 7.2 mm (95% CI: 6.5-7.8 mm) and 4.4 mm (95% CI: 4.1-4.7 mm), respectively (**Table 2**). The pooled proportions of aneurysm location are listed in **Table 3**, whereas the pooled follow-up time was 13.0 months (95% CI: 10.2-15.8 months).

**Table 2.**
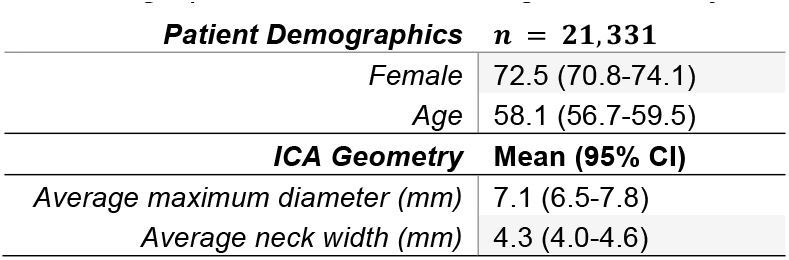
Patient demographics and meta-averages of aneurysm dimensions.

**Table 3.**
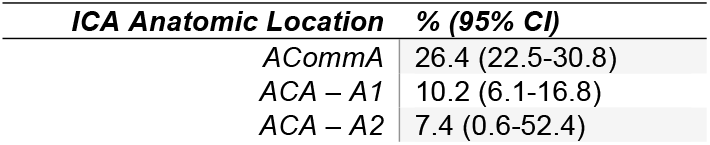
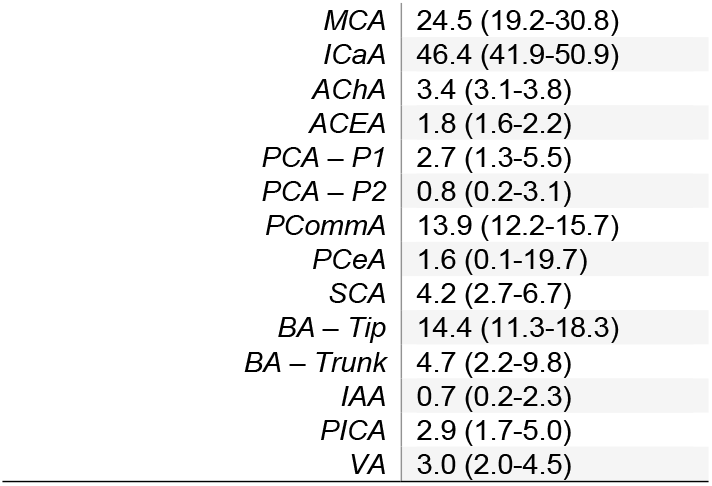
Meta-proportions of ICA anatomic location on the circle of Willis.

Overall, 20,874 ICAs had angiogram-determined occlusion rates reported immediately after treatment or at the earliest follow-up (for flow-diverters only). From this subset, 15,713 (74.9%) were treated with coiling alone, 3,206 (15.3%) with stent-assisted coiling (SAC), 782 (3.72%) with balloon-assisted coiling (BAC), 896 (4.26%) with flow diverters, 270 (1.2%) with the Woven-EndoBridge (WEB), and 158 (0.75%) with other endovascular technologies (i.e., hydrogel-coated coils, Contour Neurovascular system, and Medina).

### Occlusion Effectiveness

The meta-analysis of aneurysm occlusion effectiveness was performed in three parts: (i) the pooled meta-proportion of the immediate complete IAC occlusion (i.e., RROC-I rates), (ii) the long-term occlusion effectiveness using a LGLMM, and (iii) a comparison of the last available follow-up complete occlusion meta-proportion and the predicted probability of complete occlusion at the average follow-up time based on the LGLMM. All the analyses were performed with the extracted RROC-I rates for Coils (grouped as coiling-alone and assisted-coiling techniques), WEB device, and flow diverters. Other devices (hydrogel-coated coils, Contour Neurovascular system) were excluded from the analysis due to a relatively low number of studies available but are included in the overall discussion.

### Immediate Occlusion after Endovascular Treatment

The proportions for RROC-I rates immediately after the endovascular therapy were obtained for each study. A proportion meta-analysis was performed to pool study proportions using a random effects model. A meta-proportion for each individual device is shown in **Table 4**. These proportions represent the global complete occlusion rate obtained in all the studies for each device and its confidence interval. Flow diverters were not analyzed immediately after treatment due to the nature of its mechanism of action, where the target aneurysms become progressively occluded over time (i.e., no occlusion occurs immediately after treatment but 2-6 months after the treatment).

**Table 4.**
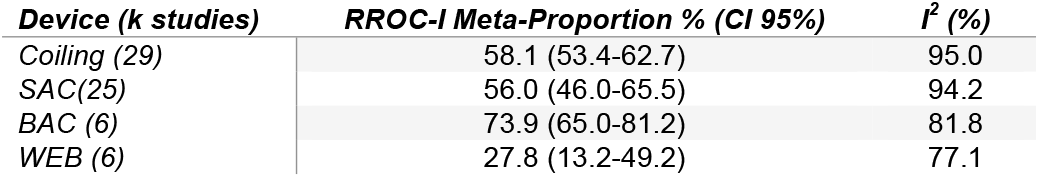
Pooled meta-proportions and heterogeneity of the RROC-I for modern endovascular devices, except flow diverters.

### Long-Term Occlusion Effectiveness of Endovascular Treatment

Binomial LGLMM models were fitted to the occlusion effectiveness data obtained from the 80 studies. Three models were obtained for the probability of RROC-I for each individual device categories as a function of angiographic follow-up time: (i) all coiling techniques (i.e., bare coils, SAC, and BAC), (ii) the WEB device, and (iii) flow diverters. The models for other devices were not performed due to a relatively low number of available studies. An individual model for BAC was attempted; however, this was not possible because of the low availability of follow-up time points for this technique. Therefore, only one model for all available coiling data was performed. The corresponding model parameters are summarized in **Table 5** and the models fits are shown in **Figure 2**. In addition, the traditional fixed-effects logistic models (FELM) were compared to the LGLMM to demonstrate how the random effects were induced by individual study variation.

**Table 5.** Model parameters for the LGLMM and FELM models of probability of RROC-I as a function of follow-up (FU) time for endovascular therapy techniques. OR: odds ratio.

| *Device* | *Parameter* | *LGLMM* |  |  | *FELM* |  |  |
| --- | --- | --- | --- | --- | --- | --- | --- |
|  |  | **OR** | **CI (95%)** | **p-value** | **OR** | **CI** | **p-value** |
| Coiling | Intercept | 2.50 | 1.51-4.15 | <0.001 | 1.15 | 1.12-1.19 | <0.001 |
|  | FU time | 1.04 | 0.98-1.09 | 0.199 | 1.03 | 1.03-1.04 | <0.001 |
| WEB | Intercept | 0.37 | 0.08-1.63 | 0.187 | 0.49 | 0.37–0.64 | <0.001 |
|  | FU time | 1.20 | 0.98-1.47 | 0.074 | 1.16 | 1.12–1.21 | <0.001 |
| Flow Diverters | Intercept | 2.63 | 0.67–10.32 | 0.165 | 0.77 | 0.65–0.91 | 0.003 |
|  | FU time | 1.05 | 0.92–1.19 | 0.452 | 1.09 | 1.07–1.10 | <0.001 |

**Figure 2.**
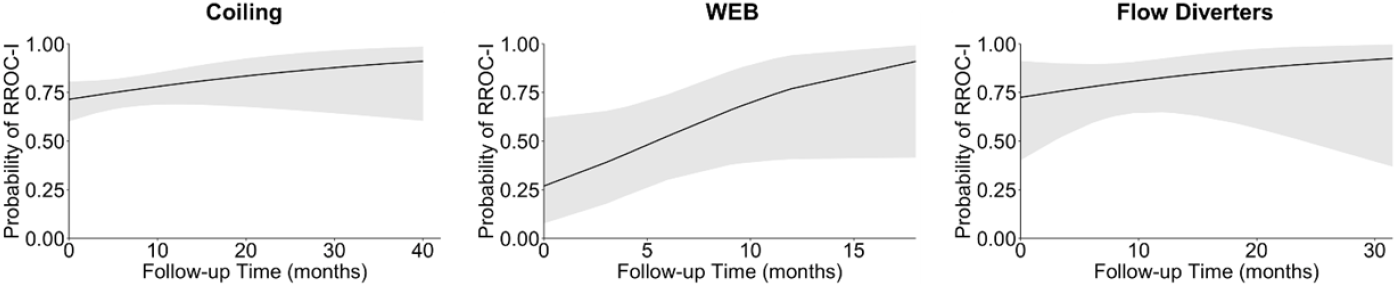
LGLMM model fits for probability of RROC-I as a funcion of follow-up time for the evaluated endovascular therapy techniques. Grey shading represents CI 95%.

Further, the pooled meta-proportions exhibit a higher degree of immediate occlusion for BAC than the other coiling techniques, whereas the WEB exhibited poor immediate occlusion degrees immediately after treatment. On the other hand, the fitted models in **Figure 2** demonstrate similar complete occlusion behaviors across the different endovascular therapy techniques. The WEB device exhibits a unique behavior, where the initial (FU time = 0 months) occlusion degree approximates the proportion described in **Table 4**, which then increases to similar levels of complete occlusion of coiling and flow diverters. Using the discrete predicted values of the models, we observed that the predicted probability of complete occlusion at 6 months for coiling, WEB, and flow diverters was 0.76 (95% CI: 0.67-0.82), 0.53 (95% CI: 0.30-0.74), and 0.78 (95% CI: 0.60-0.90). At 11-12 months, the probability increased to 0.79 (95% CI: 0.69-0.86) for coils, 0.74 (0.40-0.92) for WEB, and 0.83 (0.65-0.92) for flow diverters. Finally, at the last available FU point RROC-I probability was 0.91 (95% CI: 0.60-0.99) at 40 months for coiling, 0.91 (95% CI: 0.41-0.99) at 18 months for WEB, and 0.92 (95% CI: 0.37-1.00) for flow diverters. Overall, long-term RROC-I probability was homogeneous across endovascular techniques.

## DISCUSSION

### Overall Remarks and Occlusion Effectiveness

In this study, 19,751 aneurysms were treated with coil-based techniques in the 22-year span of the included studies. However, coiling exhibited limitations in its early years related to procedural complications, such as aneurysm/parent vessel perforation, mass effect, parent vessel occlusion and/or coil migration.^21^ In addition, the long-term efficacy of coiling is limited by potential coil compaction and/or aneurysm recurrence, thus leading to the need for retreatment of the aneurysm. These limitations led to the development of assisting techniques for coil embolization. For example, the BAC emerged as a safe alternative for ICAs with unfavorable sack-to-neck ratios, and the SEC for more complex geometries (giant, fusiform) with difficult-to-treat locations, such as bifurcation ICAs. Nonetheless, the overall coiling techniques suffer from limitations, including aneurysm recurrence, difficulty to treat aneurysms with tortuous parent vessels, and high re-treatment rates.^22^

Based on the limitations of endovascular coil embolization, flow diverters emerged as a great alternative for ICA treatment. Flow diverters are similar to stents in terms of fabrication methods and materials (both are tubular wire meshes) but differ in porosity and flexibility. They aim to decouple blood flow between the parent vessel and the aneurysm space, allowing progressive thrombus formation within the aneurysm sac. In addition, the wired mesh acts as a scaffold for neo-endothelization, which can lead to complete parent vessel remodeling.^23^ This approach has expanded the application of endovascular therapy to ICA geometries and locations that are not treatable with coiling techniques. Finally, endosaccular flow diverters, like the WEB, have expanded flow diverting technology to bifurcation aneurysms.

In this study, we aimed to understand the potential of these techniques for the treatment of saccular unruptured aneurysms, a broad ICA category that can be treated with most endovascular approaches. Due to the greater effect of the immediate complete occlusion in recurrence rates and retreatment,^16,24^ understanding the overall RROC-I rate of the most prominent endovascular therapies is of paramount importance for improved therapeutic selection and the development of novel, individualized endovascular technologies.

The results of this meta-analysis from 22,193 unruptured saccular ICAs showed several key differences and similarities between endovascular devices. We found that the immediate ICA occlusion is improved in the BAC, when compared to other coiling techniques (73.8% [95% CI: 65.0%-81.1%] vs 56.7% [95% CI: 52.5%-60.8%]), as described previously in the literature. These differences contribute to the debate on the differences between coiling techniques, where occlusion rates have been found to be similar for coiling alone, SAC and BAC.^25-27^ In addition, our results validate previous results for RROC-I rates immediately after treatment; we found that RROC-I pooled proportion was 27.7% (95% CI: 13.2%-49.2%), similar to what Asnafi *et al*. found in their meta-analysis for the same device: 27.0% (95% CI: 15.0%-39.0%).^28^

In the longer-term post aneurysm embolization, our results indicated that different endovascular therapies provide similar levels of RROC-I rates. In the last available follow-up point, coiling, the WEB, and flow diverters exhibited >90% occlusion (**Figure 2**). However, the WEB device reached these levels of occlusion in a much shorter time (18 months), while the coiling techniques and flow diverters required at least 40 and 31.5 months, respectively, to achieve the same probability of RROC-I. Quantitatively, this was reflected by a greater odds ratio for the WEB in the GLMM model (**Table 5**). These findings would be useful to inform current guidelines for method selection for ICA endovascular therapy.

### Occlusion Efficacy of Other Devices

In this study, we focused on obtaining the comprehensive information about modern endovascular devices used in studies that fulfilled our I/E criteria. However, not all the included studies could be analyzed in a meta-analysis (e.g., due to a low number of available studies). Specifically, two devices from two separate studies that fulfilled our I/E criteria were not included in this meta-analysis. The devices that were not analyzed were hydrogel-coated coils (HCCs) and the Contour Neurovascular System (CNS).

The CNS is an intrasaccular flow diverter with a similar mechanism of action as the WEB device. However, their geometries differ from the WEB and make the CNS a separate field of study for this analysis. The CNS is a braided wire mesh that is designed specifically for wide-necked bifurcation aneurysms. It has an “upside-down-umbrella” geometry that resembles the original geometry of a bifurcation when deployed in the aneurysm ^29^. The included study by Biondi *et al*. (2022) ^14^is a retrospective single-arm study where the CNS was tested in 53 patients. Complete occlusion using the CNS was 16.7% immediately after treatment, 79.6% at 3 months follow-up, and 51.8% at 1 year follow-up. It is not common to see a reduction in complete occlusion when data is available for the whole cohort across follow-up times, and this reduction is not explained in the study discussion. Future research for the CNS should shed more light on the behavior observed in this study. Also, the authors indicate that the patient cohort included their “learning curve” period for the CNS, which can also yield to biased results. Overall, the CNS is a promising device that requires further and prospective research to be included in systematic meta-analysis studies.

On the other hand, the HCCs are an advancement of coil-based technology where the coil is coated with a polymeric sheath, which absorbs water and swells to occupy gaps between coils. This technology was developed to increase packing density of traditional coils. Several trials have been performed to study the advantage of the HCCs over traditional coiling, but no significant differences have been found in immediate complete occlusion rates ^30,31^. In our meta-analysis, only one study pertinent to the use of the HCCs was accepted for inclusion. Poncyljusz *et al*. ^32^ performed a single-center controlled randomized trial to compare bare platinum coils (BPCs) and the HCCs, but reported no significant difference in occlusion effectiveness. The HCCs data from this study was not included in any of the meta-analyses due to the only data point for the ICA therapy with HCCs.

### Implications for the Development and Translation of Endovascular Devices

Currently, most novel endovascular devices are compared with the GDCs in their pre-clinical and clinical trials. However, with the growth of flow diverting technology (both intra-saccular and extra-saccular), we questioned the *gold standard* status of the GDCs for unruptured aneurysms. Our results demonstrated that immediate occlusion of unruptured saccular ICAs is homogeneous across different coiling technologies, and that other endovascular devices do not offer an advantage over the use of the GDCs for unruptured saccular ICAs. However, the long-term complete occlusion of ICAs is more likely to occur after 18 months of treatment when the WEB devices are used. These findings could guide the clinical decisions on selecting an endovascular device that is better suited for the effectiveness evaluation of novel endovascular embolization system for unruptured ICA therapy.

### Study Limitations

A meta-analysis is understood as the highest form of scientific evidence. However, most meta-analyses suffer from a high degree of heterogeneity between the studies used to perform them, in addition to very low availability of data that suffices I/E criteria in the literature. These factors played a significant role in our study. This heterogeneity can be induced by differences in study design, prospective or retrospective data generation and gathering, experience of neurosurgeons with the studied endovascular device, use of internal or centralized assessment of angiographic outcomes, among several other factors. In this study, we aimed at reducing the chances of Type II error by including study-specific variability in the assessment of our data. To do this, all our meta-analyzed metrics (averages and proportions) were calculated using random effects models, which have the great advantage of providing homogeneous weights for all studies in the analysis. In addition, the long-term modeling of RROC-I probability as a function of follow-up time in months was performed using a logistic GLMM, which also accounts for the random effects associated with study-specific variability.

Another important limitation in our analysis is related to a low availability of angiographic outcomes in the long-term follow-up (>2 years) of ICAs treated with endovascular devices. This is related to different factors: (i) the WEB became available for clinical use in Europe in 2011 and later in other regions; long-term prospective trials have started to become available recently, but more data is expected to be published in the coming decade; (ii) flow diverters can be applied to more ICA geometries than other endovascular devices, including fusiform and dissecting aneurysms; in this study, we aimed at including unruptured saccular aneurysms only, which led to the rejection of a relatively high number of studies that reported angiographic outcomes for all types of geometries together; (iii) we intended to perform analysis of the HCCs as individual endovascular devices; however, it is frequently applied as a “finishing” coil after occupying most of the aneurysm space with bare GDCs. The reporting of this type of procedure is inconsistent across literature, which led our protocol to be designed to only include studies where the HCCs were used as the only type of coil in the aneurysm; (iv) novel devices, including the Medina embolic device,^33^ the CNS, the LUNA embolization system^34^, among other flow diverting devices are still “young” in the endovascular market, and their available data did not fulfill our I/E criteria.

## Conclusions

Endovascular therapy for unruptured saccular ICAs has benefited from the emergence of novel technology in the last decade. Intra-saccular and extra-saccular flow diverters are some of the most notable technologies for the treatment of a wide array of ICA geometries, rupture status, and anatomic location. However, the GDCs, and the assisting technology used to improve its applicability, remains a great tool for endovascular embolization of unruptured saccular ICAs. The immediate RROC rates for the GDCs are yet to be improved and future developments in endovascular ICA therapeutics should aim to address this long-standing limitation to reduce aneurysm recurrence. Nevertheless, the WEB device shows great promise in reducing periods for progressive complete occlusion, as demonstrated in this meta-analysis.

## Data Availability

The data will be made available upon request.

## AVAILABILITY OF DATA

Data not included in the Supplemental Data will be available upon request to the authors, including templates for data collection, a list of rejected studies after screening, raw extracted data from the included studies and scripts from R Studio for data analysis.

## AUTHOR CONTRIBUTION

S.A.P.-C. contributed to conceptualization, study design, screening of studies at all stages of the systematic search, data extraction, statistical design, and analysis, and writing the original draft. E.R.J contributed as a reviewer in the screening of studies for the systematic search and data extraction. K.A.L, L.R.T. and T.L.C. assisted in the screening of studies for the systematic search. Y.D.Z. contributed to study design and statistical analysis. B.N.B. contributed to conceptualization and manuscript editing. C.-H.L. contributed to conceptualization, supervision, manuscript editing, and funding acquisition. All authors read and approved the final article.

## REFERENCES

1. Guglielmi, G., Viñuela, F., Dion, J., et al. Electrothrombosis of saccular aneurysms via endovascular approach: Part 2: Preliminary clinical experience. Journal of Neurosurgery 75, 8, 1991. doi: 10.3171/jns.1991.75.1.0008

2. Guglielmi, G., Vinuela, F., Sepetka, I., et al. Electrothrombosis of saccular aneurysms via endovascular approach. Part 1: Electrochemical basis, technique, and experimental results. Journal of Neurosurgery 75, 1, 1991. doi: 10.3171/jns.1991.75.1.0001

3. Claiborne, J.S., Wilson, C.B., Halbach, V.V., et al. Endovascular and surgical treatment of unruptured cerebral aneurysms: Comparison of risks. Annals of Neurology 48, 11, 2000. doi: 10.1002/1531-8249(200007)48:1<11::aid-ana4>3.3.co;2-m

4. Molyneux, A.J., Kerr, R.S., Yu, L.-M., et al. International subarachnoid aneurysm trial (isat) of neurosurgical clipping versus endovascular coiling in 2143 patients with ruptured intracranial aneurysms: A randomised comparison of effects on survival, dependency, seizures, rebleeding, subgroups, and aneurysm occlusion. Lancet 366, 809, 2005. doi: 10.1053/jscd.2002.130390

5. Kang, X.K., Guo, S.F., Lei, Y., et al. Endovascular coiling versus surgical clipping for the treatment of unruptured cerebral aneurysms: Direct comparison of procedure-related complications. Medicine (Baltimore) 99, e19654, 2020. doi: 10.1097/md.0000000000019654

6. Chai, C.L., Jeon, J.P., Tsai, Y.-H., et al. Endovascular intervention versus surgery in ruptured intracranial aneurysms in equipoise. Stroke 51, 1703, 2020. doi: doi:10.1161/STROKEAHA.120.028798

7. Laurent, D., Lucke-Wold, B., Leary, O., et al. The evolution of endovascular therapy for intracranial aneurysms: Historical perspective and next frontiers. Neuroscience Insights 17, 26331055221117560, 2022. doi: 10.1177/26331055221117560

8. White, P.M., Lewis, S.C., Gholkar, A., et al. Hydrogel-coated coils versus bare platinum coils for the endovascular treatment of intracranial aneurysms (HELPS): A randomised controlled trial. Lancet 377, 1655, 2011. doi: 10.1016/S0140-6736(11)60408-X

9. Alain, B., Marta Aguilar, P., Hans, H., et al. Diversion-p64: Results from an international, prospective, multicenter, single-arm post-market study to assess the safety and effectiveness of the p64 flow modulation device. Journal of NeuroInterventional Surgery 14, 898, 2022. doi: 10.1136/neurintsurg-2021-017809

10. Tibor, B., David, F.K., Isil, S., et al. Pipeline for uncoilable or failed aneurysms: Results from a multicenter clinical trial. Radiology, 2013. doi: 10.1148/radiol.13120099

11. Piano, M., Valvassori, L., Lozupone, E., et al. Fred Italian registry: A multicenter experience with the flow re-direction endoluminal device for intracranial aneurysms. Journal of Neurosurgery, 1, 2019. doi: 10.3171/2019.1.JNS183005

12. Issa, R., Al-Homedi, Z., Syed, D.H., et al. Surpass evolve flow diverter for the treatment of intracranial aneurysm: A systematic review. Brain Sciences 12, 810, 2022. doi: 10.3390/brainsci12060810

13. van Rooij, S., Sprengers, M., Peluso, J., et al. A systematic review and meta-analysis of woven endobridge single layer for treatment of intracranial aneurysms. Interventional Neuroradiology 26, 455, 2020. doi: 10.1177/1591019920904421

14. Biondi, A., Primikiris, P., Vitale, G., et al. Endosaccular flow disruption with the contour neurovascular system: Angiographic and clinical results in a single-center study of 60 unruptured intracranial aneurysms. Journal of NeuroInterventional Surgery, 2022. doi: 10.1136/jnis-2022-019271

15. Pineda-Castillo, S.A., Stiles, A.M., Bohnstedt, B.N., et al. Shape memory polymer-based endovascular devices: Design criteria and future perspective. Polymers 14, 2526, 2022. doi: 10.3390/polym14132526

16. Sadato, A., Hayakawa, M., Adachi, K., et al. Large residual volume, not low packing density, is the most influential risk factor for recanalization after coil embolization of cerebral aneurysms. PLoS ONE 11, e0155062, 2016. doi: 10.1371/journal.pone.0155062

17. Tawfik, G.M., Dila, K.A.S., Mohamed, M.Y.F., et al. A step by step guide for conducting a systematic review and meta-analysis with simulation data. Tropical Medicine and Health 47, 46, 2019. doi: 10.1186/s41182-019-0165-6

18. Moher, D., Liberati, A., Tetzlaff, J., et al. Preferred reporting items for systematic reviews and meta-analyses: The PRISMA statement. Journal of Clinical Epidemiology 62, 1006, 2009. doi: 10.1016/j.jclinepi.2009.06.005

19. McKenzie, J.E., Brennan, S.E., Ryan, R.E., et al. Defining the criteria for including studies and how they will be grouped for the synthesis. Cochrane Handbook for Systematic Reviews of Interventions2019. pp. 33. doi: 10.1002/9781119536604.ch3

20. Balduzzi, S., Rücker, G., and Schwarzer, G. How to perform a meta-analysis with R: A practical tutorial. Evidence Based Mental Health 22, 153, 2019. doi: 10.1136/ebmental-2019-300117

21. Murayama, Y., Nien, Y.L., Duckwiler, G., et al. Guglielmi detachable coil embolization of cerebral aneurysms: 11 years’ experience. Journal of Neurosurgery 98, 959, 2003. doi: 10.3171/jns.2003.98.5.0959

22. Lee, K.S., Zhang, J.J.Y., Nguyen, V., et al. The evolution of intracranial aneurysm treatment techniques and future directions. Neurosurgical Review 45, 1, 2022. doi: 10.1007/s10143-021-01543-z

23. Ravindran, K., Salem, M.M., Alturki, A.Y., et al. Endothelialization following flow diversion for intracranial aneurysms: A systematic review. American Journal of Neuroradiology 40, 295, 2019. doi: 10.3174/ajnr.A5955

24. Grunwald, I.Q., Papanagiotou, P., Struffert, T., et al. Recanalization after endovascular treatment of intracerebral aneurysms. Neuroradiology 49, 41, 2007. doi: 10.1007/s00234-006-0153-5

25. Vivanco-Suarez, J., Wallace, A.N., Dandapat, S., et al. Stent-assisted coiling versus balloon-assisted coiling for the treatment of ruptured wide-necked aneurysms: A 2-center experience. Stroke: Vascular and Interventional Neurology 3, e000456, 2023. doi: doi:10.1161/SVIN.122.000456

26. Waldeck, S., Chapot, R., von Falck, C., et al. A comparative evaluation of standard and balloon-assisted coiling of intracranial aneurysms based on neurophysiological monitoring. Journal of Clinical Medicine 11, 677, 2022. doi: 10.3390/jcm11030677

27. Chalouhi, N., Starke, R.M., Koltz, M.T., et al. Stent-assisted coiling versus balloon remodeling of wide-neck aneurysms: Comparison of angiographic outcomes. American Journal of Neuroradiology 34, 1987, 2013. doi: 10.3174/ajnr.A3538

28. Asnafi, S., Rouchaud, A., Pierot, L., et al. Efficacy and safety of the woven endobridge (WEB) device for the treatment of intracranial aneurysms: A systematic review and metaanalysis. American Journal of Neuroradiology 37, 2287, 2016. doi: 10.3174/ajnr.A4900

29. Akhunbay-Fudge, C.Y., Deniz, K., Tyagi, A.K., et al. Endovascular treatment of widenecked intracranial aneurysms using the novel contour neurovascular system: A singlecenter safety and feasibility study. Journal of NeuroInterventional Surgery 12, 987, 2020. doi: 10.1136/neurintsurg-2019-015628

30. Xue, T., Chen, Z., Lin, W., et al. Hydrogel coils versus bare platinum coils for the endovascular treatment of intracranial aneurysms: A meta-analysis of randomized controlled trials. BMC Neurology 18, p2018. doi: 10.1186/s12883-018-1171-8

31. Taschner, C.A., Chapot, R., Costalat, V., et al. Second-generation hydrogel coils for the endovascular treatment of intracranial aneurysms: A randomized controlled trial. Stroke 49, 667, 2018. doi: 10.1161/STROKEAHA.117.018707

32. Poncyljusz, W., Zarzycki, A., Zwarzany, Ł., et al. Bare platinum coils vs. Hydrocoil in the treatment of unruptured intracranial aneurysms: A single center randomized controlled study. European Journal of Radiology 84, 261, 2015. doi: 10.1016/j.ejrad.2014.11.002

33. Aguilar Perez, M., Bhogal, P., Martinez Moreno, R., et al. The medina embolic device: Early clinical experience from a single center. Journal of Neurointerventional Surgery 9, 77, 2017. doi: 10.1136/neurintsurg-2016-012539

34. Piotin, M., Biondi, A., Sourour, N., et al. The luna aneurysm embolization system for intracranial aneurysm treatment: Short-term, mid-term and long-term clinical and angiographic results. Journal of Neurointerventional Surgery 10, e34, 2018. doi: 10.1136/neurintsurg-2018-013767

