## Supplemental Tables 1-4 for "Systematic Review and Meta-Analysis of Endovascular Therapy Effectiveness for Unruptured Saccular Intracranial Aneurysms"

### SUPPLEMENTAL DATA

Sergio A. Pineda-Castillo, Evan R. Jones, Keely A. Laurence, Lauren R. Thoendel, Tanner L. Cabaniss, Yan D. Zhao, Bradley N. Bohnstedt, and Chung-Hao Lee[4*]

[*] Corresponding Author

Contact Information (author name, e-mail, phone number, full mailing address):

C.-H. Lee,, +1 (405)325-4842, 865 Asp Ave. Felgar Hall 219C, 865 Asp Ave., Norman, OK 73019, USA

**S1 –** List of queries used for the systematic search of studies assessing the efficacy of endovascular devices for the occlusion of unruptured saccular intracranial aneurysms.

Database search will be performed in PubMed first with the following search conditions:

1. ("aneurysm"[TiAb] OR “intracranial aneurysm”[MeSH] OR "aneurysms"[TiAb])
2. AND ("unruptured"[TiAb] OR "incidental"[TiAb])
3. AND ("endovascular"[TiAb] OR "surgically "[TiAb] OR "surgical"[TiAb] OR "treatment"[TiAb] OR "treated"[TiAb] OR "management"[TiAb] OR "repair"[TiAb] OR "coiling"[TiAb] OR “endovascular procedure”[MeSH] OR “neurosurgical procedure”[MeSH] OR "coil"[TiAb] OR "coils"[TiAb] OR "flow-diverter"[TiAb] OR "embolization"[TiAb] OR "occlusion"[TiAb] OR "surgery"[TiAb] OR "preventive"[TiAb] OR "prevention"[TiAb] OR "procedure"[TiAb] OR "hydrogel"[TiAb] OR "liquid"[TiAb] OR "liquid-embolic"[TiAb] OR "WEB"[TiAb] OR "Woven EndoBridge"[TiAb] OR "GDC"[TiAb])
4. AND ("risk"[TiAb] OR "outcome"[TiAb] OR "Raymond-Roy"[TiAb] OR "degree"[TiAb])
5. Then, search will be performed in Cochrane with the following search conditions:
6. ("aneurysm" OR “intracranial aneurysm” OR "aneurysms") AND ("unruptured" OR "incidental") AND ("endovascular" OR "surgically " OR "surgical" OR "treatment" OR "treated" OR "management" OR "repair" OR "coiling" OR “endovascular procedure” OR “neurosurgical procedure” OR "coil" OR "coils" OR "flow-diverter" OR "embolization" OR "occlusion" OR "surgery" OR "preventive" OR "prevention" OR "procedure" OR "hydrogel" OR "liquid" OR "liquid-embolic" OR "WEB" OR "Woven EndoBridge" OR "GDC") AND ("risk" OR "outcome" OR "Raymond-Roy" OR "degree").

This conditional query covers all applicable key words we expect to observe in the selected studies.

Then, search will be complemented with 3 additional databases:

1. Cochrane.
2. ClinicalTrials.gov.
3. Web of Science.

**S2 –** Inclusion/Exclusion criteria for the screening of studies prior to data extraction.

*Inclusion:*

1. Date range: 2000-2022.
2. Clinical trials testing the occlusion effectiveness of endovascular devices (concentric coils, flow diverters, hydrogel-coated coils, WEB device, balloons, liquid embolic device) for the first line of treatment for unruptured intracranial saccular aneurysms.
3. At least 10 patients of 18 years of age and older.
4. Report of rupture status of treated aneurysm.
5. Report of risk factors (smoking habits, hypertension).
6. Report of region (Americas, Asia, Europe, Africa, etc.).
7. Report of patient demographics (sex, age, etc.).
8. Report of aneurysms characteristics (size, shape, location, parent artery).
9. Report of a post-operatory follow-up (longitudinal designs) reporting occlusion degree, fatality, reoperation.
10. Report of recurrence rates.
11. Include crude or adjusted effect estimates with corresponding 95% CIs or mean +/- SD for risk factors of clinical complications available or retrievable from the data.

*Exclusion:*

1. Language different to: English, Spanish, Italian, French, German, Dutch, Portuguese.
2. Duplicates.
3. Unrelated articles.
4. Abstract-only OR clinical protocol papers.
5. Case studies.
6. Unavailable papers.
7. Animal studies.
8. Studies where the occlusion degree of aneurysms is not measured OR it is measured with classifications that cannot be translated to RROC.
9. Studies where target aneurysms are trauma-induced, or pseudoaneurysms.
10. Studies where target aneurysms are mycotic or pathogen-induced aneurysms.
11. Studies where target aneurysms are of fusiform, lobular (sac within main sac), dissecting OR other non-saccular geometries.
12. Studies with target aneurysms associated with arterial malformations (such as collagen disorders, Moyamoya disease or syndrome, dwarfism, or autoimmune disorders).
13. Studies where the reported outcome (RROC) is not distinguished between ruptured and unruptured aneurysms, OR the study targets ruptured-only aneurysms.

**S3 –** Complete list of studies included for data-extraction.

| First Author Last Name | Year | Journal | Title |
|---|---|---|---|
| Abdelkhalek | 2022 | Neurological Sciences | Predictors of flow diverter stent in large and giant unruptured intracranial aneurysms, single-center experience |
| Aguilar-Salinas | 2019 | Cureus | Safety and efficacy of stent-assisted coiling in the treatment of unruptured widenecked intracranial aneurysms: A single center experience |
| Al-Schameri | 2021 | Acta Neurochirurgica | Microsurgical and endovascular treatment of unruptured cerebral aneurysms by European hybrid neurosurgeons to balance surgical skills and medical staff management |
| Baek | 2019 | British Journal of Neurosurgery | Initial multicentre experience using the neuroform atlas stent for the treatment of un-ruptured saccular cerebral aneurysms |
| Bhogal | 2017 | Journal of Neurointerventional Surgery | Use of flow diverters in the treatment of unruptured saccular aneurysms of the anterior cerebral artery |
| Bhogal | 2019 | Clinical Neuroradiology | Treatment of unruptured, tandem aneurysms of the ICA with a single flow diverter |
| Bhogal | 2018 | World Neurosurgery | The use of flow diversion in vessels 2.5 mm in diameter: A single-center experience |
| Bhogal | 2018 | Clinical Neuroradiology | Management of unruptured saccular aneurysms of the M1 segment with flow diversion |
| Biondi | 2022 | Journal of Neurointerventional Surgery | Endosaccular flow disruption with the contour neurovascular system: angiographic and clinical results in a single-center study of 60 unruptured intracranial aneurysms |
| Cagnazzo | 2020 | Journal of Neurointerventional Surgery | Flow modification on the internal carotid artery bifurcation region and A1 segment after M1-internal carotid artery flow diverter deployment |
| Celik | 2022 | Nature Scientific Reports | Comparison of angiographic outcomes and complication rates of WEB embolization and coiling for treatment of unruptured basilar tip aneurysms |
| Chalouhi | 2014 | Stroke | Extending the indications of flow diversion to small, unruptured, saccular aneurysms of the anterior circulation |
| Chalouhi | 2013 | Stroke | Comparison of flow diversion and coiling in large unruptured intracranial saccular aneurysms |
| Cho | 2014 | Clinical Neuroradiology | Single-stage coil embolization of multiple intracranial aneurysms: Technical feasibility and clinical outcomes |
| Chung | 2015 | Journal of Neurosurgery | Preliminary experience with self-expanding closed-cell stent placement in small arteries less than 2 mm in diameter for the treatment of intracranial aneurysms |
| Consoli | 2014 | Journal of Neurointerventional Surgery | Assisted coiling of saccular wide-necked unruptured intracranial aneurysms: Stent versus balloon |
| D'Urso | 2012 | American Journal of Neuroradiology | Coiling for paraclinoid aneurysms: time to make way for flow diverters? |
| Dammann | 2014 | Neurosurgical Review | Outcome for unruptured middle cerebral artery aneurysm treatment: surgical and endovascular approach in a single center |
| De Leacy | 2018 | Journal of Neurointerventional Surgery | Wide-neck bifurcation aneurysms of the middle cerebral artery and basilar apex treated by endovascular techniques: A multicentre, core lab adjudicated study evaluating safety and durability of occlusion (BRANCH) |
| Di | 2020 | Journal of Clinical Neuroscience | Clinical and angiographic outcomes of stent-assisted coiling of paraclinoid aneurysms: Comparison of LVIS and Neuroform stents |
| DiMaria | 2015 | American Journal of Neuroradiology | Flow diversion versus standard endovascular techniques for the treatment of unruptured carotid-ophthalmic aneurysms |

**S3 (*continued*) –** Complete list of studies included for data-extraction.

| First Author Last Name | Year | Journal | Title |
|---|---|---|---|
| Diaz | 2012 | World Neurosurgery | Middle cerebral artery aneurysms: A single-center series comparing endovascular and surgical treatment |
| Feng | 2022 | Journal of the Chinese Medical Association | Flow-diverter stent to manage intracranial aneurysms: A single center experience |
| Fields | 2011 | Journal of Neurointerventional Surgery | Stent assisted coil embolization of unruptured middle cerebral artery aneurysms |
| Ge | 2017 | Interventional Neuroradiology | The role of endovascular treatment in unruptured basilar tip aneurysms |
| Ge | 2022 | Frontiers in Neurology | Endovascular treatment of large or giant basilar artery aneurysms using the pipeline embolization device: Complications and outcomes |
| Gerlach | 2006 | Journal of Neurology, Neurosurgery & Psychiatry | Treatment related morbidity of unruptured intracranial aneurysms: Results of a prospective single centre series with an interdisciplinary approach over a 6-year period |
| Sturiale | 2022 | Neurosurgical Review | Clipping versus coiling for treatment of middle cerebral artery aneurysms: A retrospective Italian multicenter experience |
| Suzuki | 2022 | Neurologia Medico-Chirurgica | Comparison of pipeline embolization and coil embolization for the treatment of large unruptured paraclinoid aneurysms |
| Teranishi | 2022 | Neurologia Medico-Chirurgica | Preliminary experience of the surpass streamline flow diverter for large and giant unruptured internal carotid artery aneurysms |
| Yan | 2020 | Interventional Neuroradiology | The use of single low-profile visualized intraluminal support stent-assisted coiling in the treatment of middle cerebral artery bifurcation unruptured wide-necked aneurysm |
| Gherasin | 2015 | Department of Interventional Neuroradiology | Endovascular treatment of wide-neck anterior communicating artery aneurysms using WEB-DL and WEB-SL: Short-term results in a multicenter study |
| Goertz | 2021 | Neurosurgery | Woven Endobridge embolization versus microsurgical clipping for unruptured anterior circulation aneurysms: A propensity score analysis |
| Goertz | 2019 | World Neurosurgery | Extending the indication of Woven Endobridge (WEB) embolization to internal carotid artery aneurysms: A multicenter safety and feasibility study |
| Gordham | 2011 | Neurological Research | Stent-assisted aneurysm coil embolization: Safety and efficacy at a low-volume center |
| Greve | 2021 | Clinical Neuroradiology | Initial Raymond-Roy occlusion classification but not packing density defines risk for recurrence after aneurysm coiling |
| Haeren | 2022 | World Neurosurgery | Fast transition from open surgery to endovascular treatment of unruptured anterior communicating artery aneurysms – A retrospective analysis of 128 patients |
| Heo | 2019 | Korean Neurosurgery | Selective temporary stent-assisted coil embolization for intracranial wide-necked small aneurysms using solitaire AB retrievable stent |
| Hetts | 2014 | American Society of Neuroradiology | Stent-assisted coiling versus coiling alone in unruptured intracranial aneurysms in the matrix and platinum science trial: Safety, efficacy, and mid-term outcomes |
| Huang | 2019 | World Neurosurgery | Clipping versus coiling in the management of unruptured aneurysms with multiple risk factors |
| Hwang | 2011 | Journal of Neuroradiology | Endovascular treatment for unruptured intracranial aneurysms in elderly patients: Single-center report |
| Kabbasch | 2019 | Clinical Neuroradiology | The barrel vascular reconstruction device: A retrospective, observational multicentric study |

**S3 (*continued*) –** Complete list of studies included for data-extraction.

| First Author Last Name | Year | Journal | Title |
|---|---|---|---|
| Kabbasch | 2019 | World Neurosurgery | Comparison of WEB embolization and coiling in unruptured intracranial aneurysms: Safety and efficacy based on a propensity score analysis |
| Khalid | 2019 | Acta Neurochirurgica | Efficiency and complications of Woven Endobridge (WEB) devices for treatment of larger, complex intracranial aneurysms: A single-center experience |
| Kim | 2022 | Journal of Cerebrovascular and Endovascular Neurosurgery | Y-stent-assisted coiling with Neuroform Atlas stents for wide-necked intracranial bifurcation aneurysms: A preliminary report |
| Kim | 2021 | Journal of Neuroradiology | Safety and efficacy of stent-assisted coiling of unruptured intracranial aneurysms using low-profile stents in small parent arteries |
| Kim | 2016 | British Medical Journal | Endovascular treatment of unruptured ophthalmic artery aneurysms: Clinical usefulness of the balloon occlusion test in predicting vision outcomes after coil embolization |
| Kocur | 2019 | Polish Journal of Radiology | Endovascular treatment of small (<5 mm) unruptured middle cerebral artery aneurysms |
| Kuhn | 2018 | Interventional Neuroradiology | Flow-diverter stents for endovascular management of non-fetal posterior communicating artery aneurysms-analysis on aneurysm occlusion, vessel patency, and patient outcome |
| Kuhn | 2016 | British Medical Journal | Stent-assisted coil embolization of aneurysms with small parent vessels: Safety and efficacy analysis |
| Kumar | 2019 | World Neurosurgery | A retrospective analysis of treatment outcomes of 40 incidental cavernous carotid aneurysms |
| Kunert | 2021 | Nature Portfolio | Flow-diverting devices in the treatment of unruptured ophthalmic segment aneurysms at a mean clinical follow-up of 5 years |
| Kwon | 2014 | Acta Neurochirurgica | Endovascular coil embolization of unruptured intracranial aneurysms: A Korean multicenter study |
| Lee | 2020 | Journal of Cerebrovascular and Endovascular Neurosurgery | Endovascular coil embolization for unruptured intracranial aneurysms in patients over 80 years of age |
| Liu | 2018 | Journal of Neuroradiology | Parent artery reconstruction for large or giant cerebral aneurysms using the Tubridge flow diverter: A multicenter, randomized, controlled clinical trial (PARAT) |
| Lubicz | 2013 | Journal of Neuroradiology | WEB device for endovascular treatment of wide-neck bifurcation aneurysms |
| Lylyk | 2019 | Clinical Neuroradiology | Treatment of wide-necked bifurcation aneurysms: Initial results with the pCANvas neck bridging device |
| Moon | 2020 | Journal of Cerebrovascular and Endovascular Neurosurgery | Result of coiling versus clipping of unruptured anterior communicating artery aneurysms treated by a hybrid vascular neurosurgeon |
| Oh | 2015 | Clinical Neurology | Management strategy of surgical and endovascular treatment of unruptured paraclinoid aneurysms based on the location of aneurysms |
| Oishi | 2015 | Journal of Neurointerventional Surgery | Treatment results of endosaccular coil embolization of asymptomatic unruptured intracranial aneurysms in elderly patients |
| Oishi | 2020 | Journal of Clinical Neuroscience | Stent-assisted coil embolization of unruptured middle cerebral artery aneurysms using LVIS Jr. Stents |
| Oishi | 2012 | Journal of Neuroradiology | Endovascular therapy of 500 small asymptomatic unruptured intracranial aneurysms |
| Ozpeynirci | 2019 | Acta Neurochirurgica | WEB-only treatment of ruptured and unruptured intracranial aneurysms: A retrospective analysis of 47 aneurysms |

**S3 (*continued*) –** Complete list of studies included for data-extraction.

| First Author Last Name | Year | Journal | Title |
|---|---|---|---|
| Park | 2021 | Korean Society of Radiology | Single center experience of the balloon-stent technique for the treatment of unruptured distal internal carotid artery aneurysms: Sharing a simple and reliable tip to use Scepter-Atlas combination |
| Petr | 2016 | Journal of Neuroradiology | Current trends and results of endovascular treatment of unruptured intracranial aneurysms at a single institution in the flow-diverter era |
| Petrov | 2021 | Interventional Neuroradiology | Initial experience with the novel p64mw HPC flow diverter from a cohort study in unruptured anterior circulation aneurysms under dual antiplatelet medication |
| Pierot | 2008 | American Heart Association | Immediate clinical outcome of patients harboring unruptured intracranial aneurysms treated by endovascular approach – Results of the ATENA study |
| Pierot | 2010 | American Heart Association | Endovascular treatment of very small unruptured aneurysms: Rate of procedural complications, clinical outcome, and anatomical results |
| Pierot | 2009 | Radiology | Endovascular treatment of unruptured intracranial aneurysms: Comparison of safety of remodeling technique and standard treatment with coils |
| Poncyljusz | 2020 | Journal of Clinical Medicine | Stent-assisted coiling of unruptured MCA aneurysms using the LVIS Jr. device: A multicenter registry |
| Poncyljusz | 2015 | European Journal of Radiology | Bare platinum coils vs. Hydrocoil in the treatment of unruptured intracranial aneurysms – A single center randomized controlled study |
| Pop | 2020 | Interventional Neuroradiology | Balloon-assisted coiling of intracranial aneurysms using the Eclipse 2L double lumen balloon |
| Popielscki | 2018 | Journal of Neuroradiology | Two-center experience in the endovascular treatment of ruptured and unruptured intracranial aneurysms using the WEB device: a retrospective analysis |
| Raslan | 2011 | Neurosurgery | Neuroform stent-assisted embolization of incidental anterior communicating artery aneurysms: Long-term clinical and angiographic follow-up |
| Samaniego | 2018 | Interventional Neurology | LVIS Jr device for Y-stent-assisted coil embolization of wide-neck intracranial aneurysms: A multicenter experience |
| Sato | 2021 | Turkish Neurosurgery | Comparison of stent-assisted coiling for unruptured internal carotid artery aneurysms between LVIS or LVIS Jr. and Enterprise VRD: A retrospective and single-center analysis |
| Satow | 2020 | Japan Neurosurgical Society | Coil embolization for unruptured intracranial aneurysms at the dawn of stent era: Results of the Japanese registry of neuroendovascular therapy (JR-NET) |
| Sedat | 2018 | Neuroradiology | Stent-assisted coiling of intracranial aneurysms using LEO stents: Long-term follow-up in 153 patients |
| Shigematsu | 2013 | Stroke | Endovascular therapy for asymptomatic unruptured intracranial aneurysms: JR-NET and JR-NET2 findings |
| Shimizu | 2016 | Journal of Neuroradiology | Endovascular treatment of unruptured paraclinoid aneurysms: Single-center experience with 400 cases and literature review |

**S4 –** Country distribution of the studies included in the meta-analysis.

| ***Country*** | ***Study count - n (%)*** |
|---|---|
| *Austria* | 2 (2.5) |
| *China* | 6 (7.5) |
| *Denmark* | 1 (1.2) |
| *Egypt* | 1 (1.2) |
| *Finland* | 1 (1.2) |
| *France* | 8 (10) |
| *Germany* | 15 (19) |
| *Italy* | 2 (2.5) |
| *Japan* | 10 (12) |
| *Korea* | 13 (16) |
| *Poland* | 4 (5) |
| *Russia* | 1 (1.2) |
| *Taiwan* | 1 (1.2) |
| *USA* | 14 (18) |
| *International* | 1 (1.2) |
